# Modeling hepatitis C virus kinetics during liver transplantation highlights the role of the liver in virus clearance

**DOI:** 10.1101/2020.08.03.20167387

**Authors:** Louis Shekhtman, Miquel Navasa, Natasha Sansone, Gonzalo Crespo, Gitanjali Subramanya, Tje Lin Chung, Sofía Pérez-del-Pulgar, Alan S. Perelson, Scott J. Cotler, Susan L. Uprichard, Harel Dahari, Xavier Forns

## Abstract

While the liver, specifically hepatocytes, are widely accepted as the main source for hepatitis C virus (HCV) production, the role of the liver/hepatocytes in the clearance of circulating HCV remains largely unknown. Here we evaluated the function of the liver/hepatocytes in clearing virus from the circulation by investigating viral clearance during liver transplantation and from culture medium in vitro. Frequent HCV kinetic data during liver transplantation were recorded from 5 individuals throughout the anhepatic (AH) phase and for 4 hours after reperfusion (RP), along with recordings of fluid balances. Using mathematical modeling, the serum viral clearance rate, *c*, was estimated. Analogously, we monitored the clearance rate of HCV at 37°C from culture medium in vitro in the absence and presence of chronically infected Huh7 human hepatoma cells. During the AH phase, in 3 transplant cases viral levels remained at pre-AH levels, while in the other 2 cases HCV declined (half-life, t_1/2_~1h). Immediately post-RP, virus declined in a biphasic manner in Cases 1-4 consisting of an extremely rapid (median t_1/2_=5min) decline followed by a slower decline (HCV t_1/2_=67min). In Case 5, HCV remained at the same level post-RP as at the end of AH. Declines in virus level were not explained by adjusting for dilution from IV fluid and blood products. Consistent with what was observed in the majority of patients in the anhepatic phase, the t_1/2_ of HCV in cell culture was much longer in the absence of chronically HCV-infected Huh7 cells. Therefore, kinetic and modeling results from both in vivo liver transplantation cases and in vitro cell culture studies suggest that the liver plays a major role in clearing HCV from the circulation.

## Introduction

The liver is widely accepted as the main site for hepatitis C virus (HCV) production but its role in the clearance of circulating HCV remains largely unknown. To determine the function of the liver in clearing cell-free virus from the circulation in patients requires viral kinetics during liver transplantation as only then can viral levels be measured in the blood during the anhepatic phase (AH) when the native liver has been removed and compared to after reperfusion (RP) of the new liver. Previous kinetic studies ^1-4^ have suggested that HCV was cleared at the same rate during the AH phase and early after RP. However, these estimates were based on limited data, obtained at the beginning and end of AH and several hours after RP. Interestingly, Garcia-Retortillo et al^4^ reported a patient with a prolonged AH of 20 hours in whom the HCV half-live (t_1/2_) during AH and after PR was estimated to be 10.3 hours and 3.8 hours, respectively, suggesting that viral clearance occurs relatively slowly during AH and increases after PR. Garcia-Retortillo et al^4^ hypothesized that after PR, massive entrance of HCV into the hepatocytes or HCV uptake by the liver reticuloendothelial system is the cause, at least in part, of HCV clearance. However, a detailed viral kinetic study during AH and immediately after PR has not been performed to test this hypothesis.

Here, we measured viral levels very frequently in 5 liver transplant cases during the AH phase and early during the post-RP period along with recording fluid balances during the surgical procedure. The viral clearance rate, *c*, during these two distinct phases was estimated using mathematical modeling accounting for viral kinetics and taking into account fluid balance including infusion of red blood cells, plasma, saline, and albumin, as well as blood loss. In parallel, we monitored the clearance rate of HCV from culture medium at 37°C in the absence and presence of chronically infected Huh7 human hepatoma cells. In both cases, HCV clearance was faster in the presence of the hepatocytes/liver suggesting that the liver plays a major role in clearing HCV from circulation.

### Cases 1-5

Five individuals with median age 60 years (range, 49-68) and median BMI 30 (range, 19-34) underwent liver transplantation (Supplementary Table 1). Four of five liver donors were male with median age 69 years (range, 29-73) (Supplementary Table 1). All patients received 500 mg methylprednisolone immediately before laparotomy and 500 mg during graft reperfusion. Blood samples (2 ml) were taken before liver transplantation, every 5-15 min during the AH phase and every 5 min during the first hour of RP and every 10 min until the end of surgery. Thereafter, blood was sampled every hour for the first 6 hours, every 6 hours during the first day, and daily during the first week (Supplementary Figure 1). Transfusion of blood products, saline, and albumin were recorded. The study was approved by the Ethics Committee of our Institution and all patients provided a written informed consent.

### Viral kinetics before and during the anhepatic phase

The median viral load before the AH phase was 6.04 (range 3.71-6.40) log_10_ IU/mL (Figure 1; Table 1). During the AH phase, which lasted 1.25 h to 1.90 h, virus levels were flat in three cases (Case 1, Case 2 and Case 3) while in two patients (Case 4 and Case 5) the viral slope was 7.6 and 3.7 log_10_ IU/mL per day, respectively (Supplementary Table 2).

**Figure 1:**
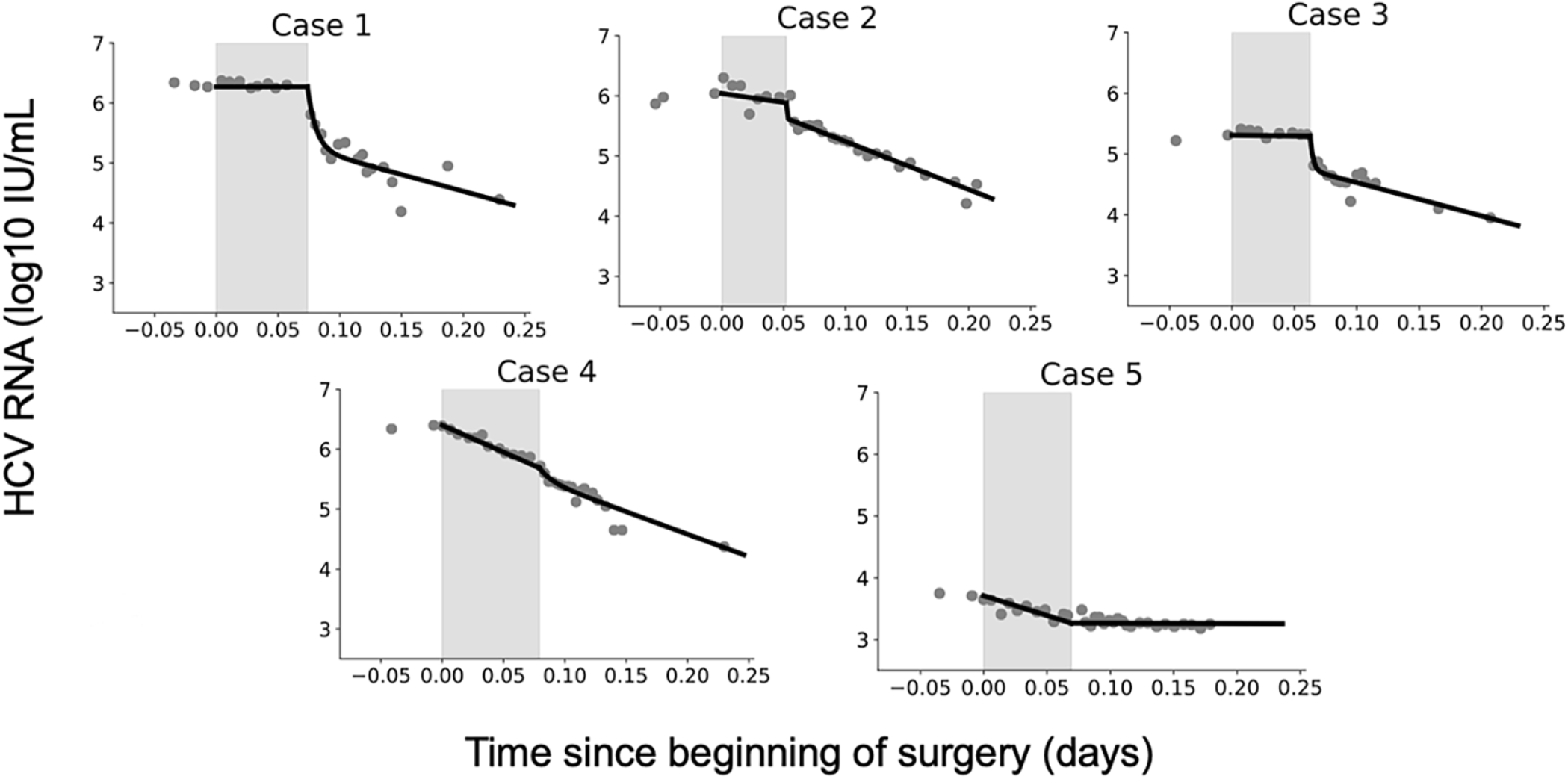
HCV RNA kinetics during liver transplantation. Serum HCV RNA kinetics in five cases before transplant, during the anhepatic phase (gray rectangles), and during the first 4 hours after liver graft reperfusion. HCV RNA measurements are shown with gray circles and model curves with solid black lines. HCV levels are graphed relevant to time of liver removal (t=0) on the x-axis. VP = a viral plateau. Model calibration and linear regression was performed using Python 3.7.4 together with Scipy Version 1.3.1 and Numpy Version 1.17.2. Results with p-values ≤ 0.05 were considered statistically significant. The viral slope was considered flat (or plateau) if the estimate was not significantly different from 0, i.e. p > 0.05.

**Table 1:** Best-fit parameter estimates of fitting Eqs. (3-4) to data obtained during the anhepatic phase, AH (clearance *c*) and 4h after graft reperfusion, RP (clearance *c*, viral load *V*_*0*_ at RP, assuming fluid volumes of 15L). The parameter κ and initial virus half-life, t_1/2_, are given only where Eq. 4 was fit (in Cases 1-4 for post RP). It is assumed that fluid intake and outtake are equal (see Supplementary File). VP, Viral Plateau (not significantly different from slope 0).

| Phase | Case | Phase Duration [days] | Num. Data Points | Total Fluid Intake [mL] | $V_0$ [log IU/mL]<sup>@</sup> | Init. HCV $t_{1/2}$* [min] | $\kappa$ [1/days] [95% CI] | HCV $t_{1/2}$ [min] [95% CI] |
| --- | --- | --- | --- | --- | --- | --- | --- | --- |
| AH | 1 | 0.074 | 8 | 0 | 6.27 | - | - | VP |
|  | 2 | 0.052 | 7 | 500 | 6.04 | - | - | VP |
|  | 3 | 0.062 | 8 | 250 | 5.31 | - | - | VP |
|  | 4 | 0.079 | 12 | 750 | 6.4 | - | - | 57 [54-60] |
|  | 5 | 0.069 | 11 | 951 | 3.71 | - | - | 79 [67-95] |
| Median (range) |  | **0.067**<br>(0.052-0.079) | **8**<br>(7-12) | **500**<br>(0-951) | **6.04**<br>(3.71-6.4) |  |  |  |
| RP | 1 | 0.17 | 15 | 0 | 6.27 | 2.9 | 143 [120-166] | 77 [53-140] |
|  | 2 | 0.17 | 22 | 1000 | 5.89 | 0.4 | 3620 [3080-4150] | 54 [50-59] |
|  | 3 | 0.17 | 15 | 250 | 5.29 | 1.9 | 399 [348, 450] | 79 [63-105] |
|  | 4 | 0.17 | 18 | 3300 | 5.69 | 15.8 | 100<sup>†</sup> | 59 [52-68] |
|  | 5 | 0.17 | 21 | 3350 | 3.27 |  | - | VP** |
| Median (range) |  | **0.17**<br>(0.17-0.17) | **18**<br>(15-22) | **1000**<br>(0-3350) | **5.69**<br>(3.27-6.27) | **5.25**<br>(0.4-15.8) | **1387**<br>(143-3620) | **67**<br>(54-79) |

\* Due to high uncertainty, the initial virus clearance rate (i.e., $c_0$ in Eq. 4) was fixed to its best-fit value (first estimated with $c_0$ , $c$ , and $\kappa$ as free parameters) and then the errors on the remaining parameters were computed.
\*\* Eq. 3 was used to estimate HCV $t_{1/2}$ .
@ For 4h after RP, the $V_0$ listed is the final value of the fit for AH. This value was then fed into the model (Eqs. 3 or 4) for calibration with HCV data 4h after RP.
†Parameter was set to this fixed value due to high uncertainty and was exempted from the calculation of median and range.

### Viral kinetics after graft reperfusion

During the first four hours after RP, HCV measurements were taken at intermittent times (Fig. 1) and volume input and output were recorded (Table 1). In Cases 1-4, HCV RNA declined in a biphasic matter, while in Case 5, HCV RNA remained suppressed at the same levels as at the end of AH phase. For Cases 1-4, the biphasic decline consisted of an initial, sharp decrease within 11-22 min, followed by a longer but slower decrease (Table 1; Supplementary Table 3). The slope of the initial decrease was 66, 36, 50 and 49 log_10_ IU/mL per day for Cases 1-4 respectively. The slower second phase slope for Cases 1-4 was 6, 8.4, and 5.3, and 8.8 log_10_ IU/mL per day, respectively (Supplementary Table 3). The viral plateau in Case 5 is distinct from the other cases, although it is worth noting that this patient’s viral load continued to be flat for ~6 days post RP (Supplementary Figure 1). The fluid input during post-RP for Cases 1-5 were 0, 1000, 250, 3300 and 3350 mLs (Table 1).

### Mathematical modeling of HCV kinetics during liver transplantation

A schematic of the mathematical modeling approach utilized to understand HCV dynamics during the different stages of liver transplantation is shown in Supplementary Figure 2. Shortly before liver removal, we assume that virus is produced at constant rate *P* and cleared from the serum at rate *c* per virion,

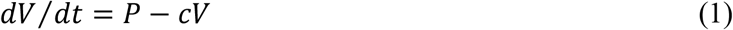

Assuming the pre-transplant viral load to be at steady-state, i.e., *dV* / *dt* = 0, implies that HCV production and clearance balance or *P* = *cV*. During the AH phase and four hours after RP, we assume that in the absence of the liver, virus production ceases, *P* ≈ 0. Hence, Equation 1 reduces to

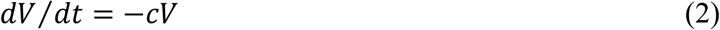

Some patients received albumin and blood transfusions to compensate for the fluid loss that occurred during transplant surgery. The blood loss causes a reduction in the absolute amount of virus in the circulation, but does not affect the virus concentration, *V*. In contrast, the fluid intake results in a dilution of *V*. We can thus describe the change in patient viral load during the AH phase as in ^3^

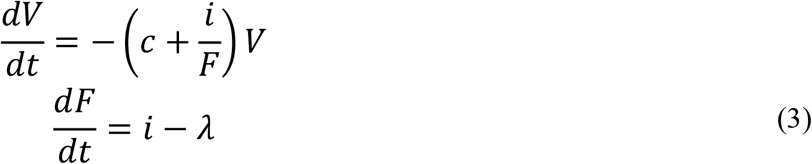

where *F* denotes the total body extracellular fluid possibly varying over the course of the AH phase, *i* is the rate of fluid intake and λ is the rate of blood/fluid loss. The total fluid volume before surgery was calculated using the patient’s body weight under the assumption of 15 L of extracellular fluid per 70 kg.

To account for the initial rapid and final slower phases of HCV decline post RP in Cases 1-4 we modified Eq. 3 by including a time-dependent viral clearance rate *c*_*t*_ (*t*):

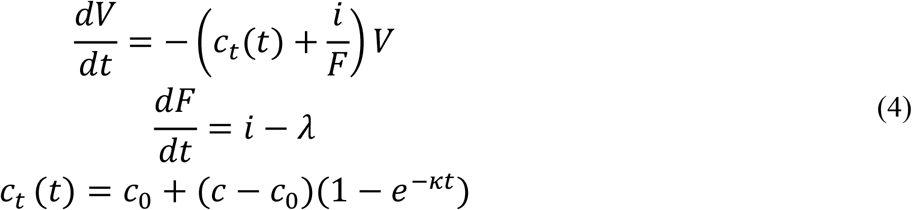

where *c*_*0*_ represents the initial rapid clearance rate, *c* the final slower clearance rate and κ governs how quickly *c*_*0*_ reduces to *c*. Note that if κ = 0 or if κ is very large then *c*_*t*_ (*t*) reduces to a constant.

The fluid input and loss rates *i,λ* were computed by the ratio of recorded volumes of fluid input and loss divided by each patient’s duration in the AH phase. In the absence of surgical complications, the fluid loss and intake are usually approximately balanced, though loss is generally slightly more than the input. Since the exact volume of blood loss was not available, we evaluated the maximum impact of fluid intake which occurs when the blood loss and input are balanced, i.e. *λ=i*. When *λ ≤ i*, the total fluid volume *F* is a non-decreasing function over time, which attains a minimum when *λ=i*. As a consequence, the impact of *i/F*, involved in describing viral concentration dynamics, is maximum if *λ=i*.

Model fits are shown in Figure 1 and viral clearance estimates are shown in Table 1. The estimated virus clearance rates during the AH phase in Cases 1-3 were ~0 (i.e., very long t_1/2_ that implies no clearance) but were significantly different from 0 in Cases 4 and 5 corresponding of HCV t_1/2_ of 1-2 hours.

During RP, in Cases 1-4, the initial viral clearance, c_0_, t_1/2_ was on the order of 5 [0.4-16] min. The second slower clearance exhibited a median time constant κ=1387/day with a t_1/2_ on the order 67 [54-79] min (Table 1 and Fig. 1). In Case 5, Eq. (3) was used because an extremely slow (or plateaued) viral load (slope of 1.4 log_10_ IU/mL/day) was observed suggesting no production and clearance during the first 6 hours post RP (Fig. 1, Table 1) or the subsequent 6 days (Supplementary Figure 1).

### HCV clearance from culture medium in vitro

To investigate the idea of HCV clearance by the liver further, we turned to the more controlled in vitro HCV infection experimental system^5^ as previously described in detail^6^ (see Cell Culture Experiment Methods in Supplementary File). To determine the effect of cells on HCV clearance in vitro, we first measured the half-life of HCV virions at 37°C in cell culture medium in the absence of cells by monitoring the decrease of encapsidated HCV RNA over time, which indicated a corresponding half-life of 108 hours (Fig. 2, left). To measure HCV half-life in cell culture medium in the presence of chronically HCV infected cells, we established a steady state chronic infection in Huh7 cells, and then inhibited the secretion of HCV from the cells using the NS5a inhibitor daclatasvir (DSV) to block HCV replication and secretion or Naringenin (NG) to block HCV secretion. In the absence of de novo virus secretion, we monitored the clearance of encapsidated HCV RNA from the culture media over time which declined with a half-life of ~14 hours (Fig. 2, right), which is 10 times faster than the clearance observed in the absence of cells. (Fig. 2, left).

**Figure 2:**
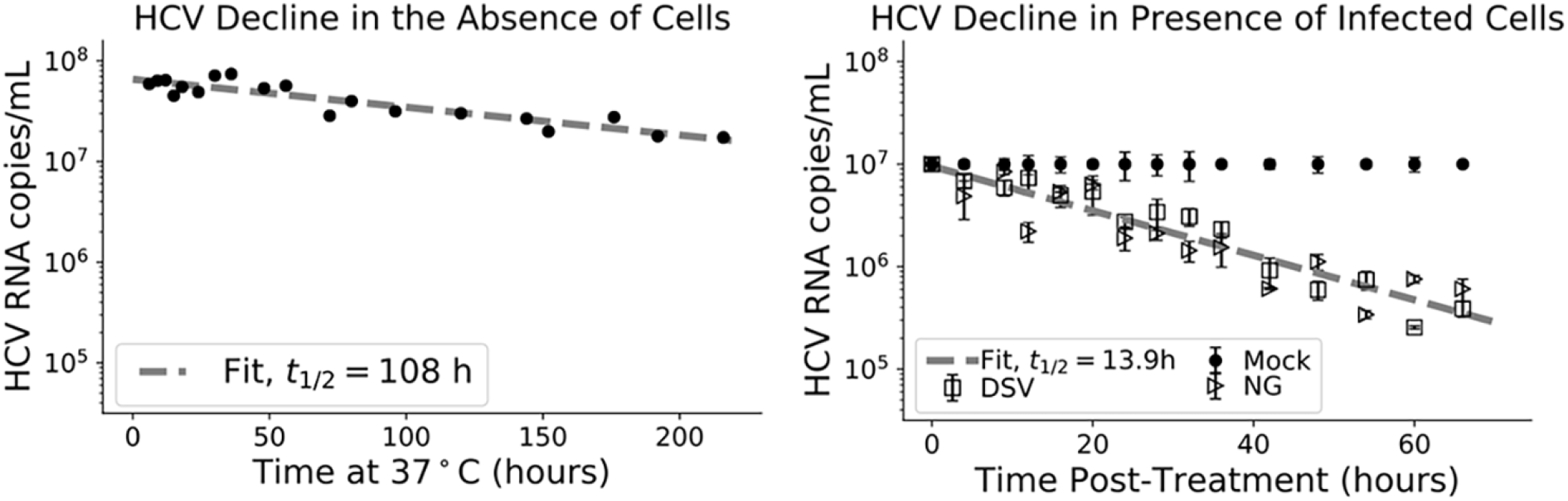
In vitro HCV RNA clearance kinetics. Left panel: Decline in encapsidated HCV RNA in culture medium at 37oC in the absence of cells. Right Panel: Decline in encapsidated HCV RNA in culture medium at 37oC in the presence of chronically infected cells. Chronically HCV infected non-growing Huh7 cells were mock treated (circles) or treated with 200μM of the HCV secretion inhibitor Naringenin (NG) (triangles)or treated with 1nM of the HCV replication and secretion inhibitor daclatasvir (DSV)(squares). In the absence of de novo HCV secretion, HCV RNA clearance from the culture medium was monitored. HCV copies were quantified by RT-qPCR and are graphed as copies/mL at the indicated times. Linear regression analysis (dashed lines) was performed to calculate virus half-life (t½).

## Discussion

The current study represented the first time that very frequent sampling for viral load monitoring during the AH phase has been performed. This frequent sampling led to the revelation that viral clearance is often minimal during the AH phase, which goes against our and others’ assumptions in previous models in which similar viral clearance during the AH and post-RP phase was assumed^1-4^. Specifically, in 3 of 5 Cases, viral load remained at a plateau in the absence of the liver suggesting not only that viral production stopped but that viral clearance stopped as well. The data suggest that the liver is involved in the clearance of circulating HCV. Interestingly, however, in Cases 4 and 5, fast viral clearance (t_1/2_~1-2 hours) was estimated via model calibration (Eq. 3) with measured data during the anhepatic phase while accounting for the recorded high fluid input of 3300 and 3350 mLs in Cases 4 and 5, respectively. Theoretically, to explain the fast viral clearance solely based on fluid input, the model (Eq. 3) predicts about 4-fold higher fluid input (i.e, ~13,000 mLs) than the recorded input (not shown), suggesting that other unknown extrahepatic mechanisms may have contributed to viral clearance during AH.

Another novel finding reported herein is that following the introduction and RP of the donor liver, the viral load declined in a biphasic manner (in Cases 1-4) consistent with an extremely fast viral decline (t_1/2_~ 5 min) that lasted ~14 min post-RP followed by slower decline (HCV t_1/2_= 67 min). We hypothesize that the initial dramatic decrease is due to the rapid entrance and binding of the virus into the new liver and/or by liver sinusoidal endothelial cells reminiscent of Ganesan et al.^7^ findings in a mouse model for adenovirus clearance 1 min after infusion. The slower decrease (i.e., HCV t_1/2_ of 67 min) represents a more physiological uptake rate of virions by the new liver and before new virions are likely to be released from infected cells. Efforts to measure HCV virion clearance have been done using antiviral drugs to inhibit viral production. Interestingly, the 67 min for HCV t_1/2_ estimated in the current study is consistent with previous estimates of the HCV t_1/2_ of 45 min in patients treated with a potent inhibitor, daclatasvir, that blocks virus production ^8^.

While in Cases 1-4, a rapid resurgence of HCV was observed 6 hours post PR, in Case 5, an extremely slow viral load slope was observed during the first 6 hours post RP (Fig. 1) and during the subsequent 6 days (Supplementary Figure 1). The lack of viral decline during the anhepatic phase and/or slow resurgence following RP in part might be explained by graft dysfunction due to ischemia-reperfusion injury altering HCV binding to its receptors on hepatocytes. Interestingly, we previously reported a slow viral load slope post RP (over 6 hours) in 3 (of 20) patients who underwent liver transplant, where 2 of the 3 patients had a large degree of ischemia-reperfusion injury^4^. In addition, closure of porto-collateral vessels is not immediate after transplantation and in some patients blood shunting might contribute to a more modest decrease in HCV viral load during RP^9^.

Cell culture experiments support the notion that hepatocytes produce and clear the virus from the extracellular space. Like in vivo, we observed that the t_1/2_ of HCV in vitro was significantly shorter in the presence of chronically infected Huh7 cells under daclatasvir or naringenin treatment. Because these well characterized drugs do not enhance the degradation rate of HCV particles in the media, the data suggest that a large amount of the virus in the media enters (or binds to) the cells.

The notion of the liver playing a significant role in the clearance of pathogens from circulation is not new. Others suggested hepatic involvement in the clearing of simian immunodeficiency virus and infused adenovirus in animals ^7, 10^. However, to our best knowledge, the current study is the first one to investigate this in vitro specifically suggesting a role for hepatocytes in the clearance of a virus, a phenomenon which may be limited to hepatotropic viruses.

The finding that the liver plays a key role in clearing HCV from the circulation has implications for clinical best practices regarding transplantation and antiviral treatment. For transplantation, the fact that viral levels remain at steady state during the AH phase, reinforces the benefit of achieving viral clearance prior to transplantation to prevent infection of the graft.

## Data Availability

Kinetic data will be available upon request.

## Acknowledgments

The study was supported, in part, by U.S. National Institute of Health grants: R01-AI078881 and R01-AI116868, Instituto de Salud Carlos III (PI15/00151 and PI13/00155) and by Secretaria d’Universitats i Recerca del Departament d’Economia i Coneixement (grant 2017_SGR_1753) and CERCA Programme/Generalitat de Catalunya, and Germany Academic Exchange Service.

Author names in bold designate shared co-first authorship

